# Digital Mental Health Service engagement changes during Covid-19 in children and young people across the UK: presenting concerns, service activity, and access by gender, ethnicity, and deprivation

**DOI:** 10.1101/2023.08.11.23293977

**Authors:** Duleeka Knipe, Santiago de Ossorno Garcia, Louisa Salhi, Lily Mainstone-Cotton, Aaron Sefi, Ann John

## Abstract

The adoption of digital health technologies accelerated during Covid-19, with concerns over the equity of access due to digital exclusion. Using data from a text-based online mental health service for children and young people we explore the impact of the pandemic on service access and presenting concerns and whether differences were observed by sociodemographic characteristics in terms of access (gender, ethnicity and deprivation). We used interrupted time-series models to assess whether there was a change in the level and rate of service use during the Covid-19 pandemic (April 2020-April 2021) compared to pre-pandemic trends (June 2019-March 2020). Routinely collected data from 61221 service users were extracted for observation, those represented half of the service population as only those with consent to share their data were used. The majority of users identified as female (74%) and White (80%), with an age range between 13 and 20 years of age,. There was evidence of a sudden increase (13%) in service access at the start of the pandemic (RR 1.13 95% CI 1.02, 1.25), followed by a reduced rate (from 25% to 21%) of engagement during the pandemic compared to pre-pandemic trends (RR 0.97 95% CI 0.95,0.98). There was a sudden increase in almost all presenting issues apart from physical complaints. There was evidence of a step increase in the number of contacts for Black/African/Caribbean/Black British (38% increase; 95% CI: 1%-90%) and White ethnic groups (14% increase; 95% CI: 2%-27%)), sudden increase in service use at the start of the pandemic for the most (58% increase; 95% CI: 1%-247%) and least (47% increase; 95% CI: 6%-204%) deprived areas. During the pandemic, contact rates decreased, and referral sources change at the start. Findings on access and service activity align with other studies observing reduced service utilization. The lack of differences in deprivation levels and ethnicity at lockdown suggests exploring equity of access to the anonymous service. The study provides unique insights into changes in digital mental health use during Covid-19 in the UK.

## 1. Introduction

The global Covid-19 pandemic has had widespread effects worldwide. Most countries, including the UK, implemented social distancing restrictions to minimize the spread of the disease and mitigate the loss of life. There is evidence of the negative effects of the Covid-19 pandemic on an economic, social^1^, and psychological level, with the lockdowns in the UK comprising of restrictive movement measures like school closures, social distancing, and lockdown restrictions limited social contact^2^. These measures have disproportionately impacted the lives of children and young people (CYP)^3–5^. The psychological impact of these public health measures has been recognised as being an important consideration during this time^6^, with an estimation of 10 million people requiring additional mental health support as a consequence of the crisis^7^.

In the UK, schools closed on March 23^rd^, 2020, and remained so until the beginning of September, although new norms and regulations for learning environments were introduced. School closures are associated with harm to the health and wellbeing of this population, with studies across twenty countries showing 18-60% of the CYP population were psychologically distressed and there was an increased drop in school attendance and access to services^8^. Experiences of the pandemic will have been especially difficult for CYP who have a history of mental/physical health problems or are from disadvantaged backgrounds. Young people accessing digital mental health services, who are already known to be more vulnerable, are at a higher probability of heightened risk, loss of progress, as well as, increased experiences of isolation and will require ongoing psychological support^9^. There is a wider need to examine changes in population health regarding digital mental health access changes, mental health concerns, and how this is affected by sociodemographic factors. Digital web-based support services were able to continue to provide consistent support to CYPs during lockdowns when social distancing meant most services had to reconfigure or pause their service provision, or some transitioned to the adoption of digital and telemedicine approaches. Some countries responded with a national digital support provision for mental health in the face of the pandemic^10^, but it is not yet clear how well these services were utilized, or whether access was equitable across sociodemographic groups.

It is also important to consider how sociodemographic characteristics, such as ethnicity, gender, and deprivation, can influence the level of need and access to support. In particular, ethnicity-related inequalities are known to be a problem for mental health access and treatment in the UK^11^. Individuals from ethnic minorities are less likely to engage in mental health face-to-face support^12^.

Racial inequalities are also exacerbated by intersection with other social determinants of health such as deprivation, gender, and other biases that mental health provisions are subject to. The online provision of mental health support has received growing interest, and varied uptake, in recent years, especially given the utility of new technologies in increasing access to services for CYP or traditionally underserved communities^13^. Online digital environments have the particular benefit of being able to overcome barriers such as distance, improving access to care, reducing waiting times, and travel costs. Its adoption is posed to be accelerated during the crisis and beyond^14^. Digital exclusion is a risk for web-based services and interventions with the potential for a more vulnerable ‘underclass’ to be developed^15^. A recent review on digital mental health highlights the careful considerations to broaden access to care with digital mental health^16^. In terms of digital exclusion knowledge, financial deprivation and mental health concerns are known factors for the maintenance and perpetuation of this exclusion^17^. As a start, it is important to explore access to these kinds of services, so we contribute reducing digital divide^18^, and find ways to prevent disparity in its technological adoption^19^.

According to studies with mental health practitioners and CYP, some of the perceived benefits of online mental health interventions are that they offer anonymity, privacy, and emotional safety due to reduced emotional proximity to the practitioner, as well as increased flexibility, control, and accessibility to treatment^20–22^. Text-based services are in a privileged position to offer this as an alternative to video consultations that will inherently breach the anonymity valued by CYP^23^. Digital mental health services may be uniquely placed as a support provider to help those who might be lacking access, are not yet at a point of crisis or are unsure if support is available during the Covid-19 pandemic.

Kooth (Kooth.com) is a digital mental health platform. The service provides a unique opportunity to examine how CYP access an online service for mental health support and how the service changed during the global crisis. We aimed to assess the impact of the pandemic on service use in terms of overall contact, referral source, type of contact, and the type of presenting issue (e.g. mental health, abuse) using interrupted time-series modeling, and explored whether the impact is different between sociodemographic groups (gender, ethnicity and deprivation level) in one digital mental health service for CYP operating in the UK. To evaluate the impact the study aimed to answer the following research questions:

i) Has there been a change in trends of contacts to the service pre- and during Covid-19 pandemic in the UK?
ii) Has there been a change in the referral source pre- and during Covid-19 pandemic in the UK?
iii) Has there been a change in service presenting concerns pre and during Covid-19 pandemic in the UK?
iv) Is there any evidence that any changes observed vary by ethnic minority status, gender, and area-level deprivation pre and during Covid-19 pandemic in the UK?

## 2. Methods

### 2.1. Study Setting

Kooth (Kooth.com) is an online text-based service for mental health and emotional wellbeing support service, available for free to CYP aged 10-25 years in the UK. The service offers free, text-based, and anonymous support with no need for a referral, alongside an ecosystem of mental health support through moderated forum boards and curated materials with moderated activities (e.g., journaling, goal-setting, community forums). Young people can interact via text-message and receive support directly with practitioners through usually 60 minutes of synchronous text-based chats through the website. Users can also have asynchronous messages with practitioners to their inbox or indirectly engage with unstructured support and engagement through a community of peers in a moderated online forum. The platform and service provision has been previously identified as a positive virtual ecosystem through its theory of change^24^. The service regularly collects and monitors access, usage information, presenting concerns, and other service user information. It is however anonymous and the personal information about the user is limited as well as the capacity to verify service users’ identity. The routinely collected data for this study was based in users who provided specific research consent to use their data within the platform.

### 2.2. Dataset

The datasets utilised in this study were routinely collected between 2019 and 2021 at one digital mental health service (Kooth.com). The dataset contained only users who consent to use their data for research purposes at registration 50.25% of the total population of service users accessing during the period accessing Kooth.com.

A total of 34 regions structured by Clinical Commissioning Groups (CCG) with unchanged resource contracts were selected from a total of 97 (N=5 decreased; N=58 increased); the selection criteria of regions was used to prevent biases due to changes in resources that may affect the demand and capacity of the online service during the pandemic (resource increase or decrease during the observational period). The study selected only regions of the service that had the same allocated number of resources before and during the pandemic, remaining therefore constant during the period.

Four datasets extracted from the service were linked and subsequently cleaned (e.g. removal of duplicates) for the analysis. The datasets contained unique user identifiers used for linkage to create one dataset containing information about access and engagement, presenting concerns during the period, user demographic information, region of access, and contact as service activity data, including logins, registration dates, and different service usage activities.

The index of multiple deprivations was added to the dataset from the Ministry of Housing, Communities and Local Government public database^25^, the CCG rank of multiple deprivations corresponding to each contract region from the service was added from the dataset, and the region of access (reported at service registration) was grouped and structured into CCGs for the service.

Presenting concerns are practitioner-reported data about the problems, issues, or difficulties that users bring to the chat sessions, community forums and other activities in the service^26^. A total of 118 reported different presenting concerns were identified for the study. Those were grouped into five high-level categories aligned with previous literature on the mental health impact of the pandemic and allowed for sufficient observations within each category to track trends over time. The service activity data was aggregated into direct and indirect interaction contacts to differentiate the online therapy provision of the service (asynchronous and synchronous chat) linked to human-to-human interaction and, online community engagement for its forums and other activities in which a human is not directly involved but will require some indirect interaction (albeit human moderation to approve content in the forums or asynchronous communication may take place). A combination of datasets and service information was uploaded to the Adolescent Data Platform (ADP) a UK secured e-research platform for analysis and collaboration, the datasets provided the group of variables used in the observational study.

### 2.3. Study Variables

Service access contacts was our outcome of interest by month. To calculate the rate of contact per user, we identified the number of active users on the platform (i.e., denominator) by counting the number of users accessing the service at least once each month between June 2019 to April 2021. The service activity is recorded in the platform engagement analytics. The service has different offerings with activities and different ways of engagement, users can engage in an online peer support forum, write in an emotional journal, request direct synchronous chats, or send asynchronous messages to practitioners. To assess whether the type of contact differed in the different periods, we categorized contacts into either direct or indirect. Direct Contacts were defined to be interactions that are human-mediated (interaction between a practitioner and user has taken place) like chats or therapeutic asynchronous messages, these are contacts for online therapy as opposed to online community forums^27^. Indirect contacts were interactions with the platform through writing content in the online community forums, setting up goals, or using an emotional journaling tool. As a digital platform, users can still interact with the service through these kinds of indirect contact interactions, despite human interaction may be required for safeguarding, safety, and moderation, it is not mediated by a human directly.

The digital service captures self-reported information about where users were referred from (educational settings; family and friends and word of mouth; primary or secondary care professionals; charitable and care organizations; and internet advertising, social media, and other sources). It also has data on the presenting issue for which a user will engage with the service. The dataset presented 118 different types of presenting concerns, which were grouped into five categories representing the main mental health impacts of the Covid-19 pandemic for CYP identified in previous literature (Supplementary Table 2).

In addition to the service engagement data above, we also utilized data collected on gender recorded at service registration (“*My gender is best described as*” : i) male; ii) female; iii) Agender; iv) Gender-fluid), and ethnicity (“*My ethnicity most closely matches*”: i) White; ii) Black/African/Caribbean/Black British; iii) Asian or Asian British; iv) Mixed/Multiple ethnic groups; and v) Not stated). Using the index of multiple deprivation of each CCG we assigned deprivation quintiles for each user based on the rank of their CCG ^25^. We used the average IMD rank of each CCG location included (out of a possible 191) to calculate deprivation quintiles (higher ranks are least deprived) for each user in relevance to their area (partially disclosed at registration and group by CCG).

### 2.4. Analysis

For this analysis we define the time of the intervention (i.e., the start of the Covid-19 pandemic) as March 2020, this coincided with the date of the first lockdown period in the UK (23 March 2020). We provide summary statistics of the sample in terms of the number of active users per month during the pre-and pandemic periods, as well as the number of contacts by type and presenting concern. Trends of the rate of contact per user (overall and by type) are presented graphically with the intervention month marked in a red-dotted line. To examine the representativeness of our sample towards the user population at Kooth.com, Chi-square comparisons and effect sizes were performed to look at distribution differences in demographic characteristics between consented users and non-consenters and the magnitude of these differences between samples using effect size calculations.

An interrupted time-series analysis was used to assess whether there was evidence of a change in online service use during the pandemic period (April 2020 – April 2021) compared to pre-pandemic (June 2019 – March 2020) trends. We pre-specified the impact model for this analysis prior to data analysis. We examined the changes in the number of monthly contacts in the pre-pandemic and during the pandemic period. We fitted Poisson regression models with a scale parameter to account for overdispersion and the number of active users as the population offset. We fitted models for the overall number of contacts and then by type and presenting concern. We also fitted models for the number of new registrations by referral source, and further stratified the overall number of contacts by sex, ethnicity, and area deprivation level. We used the ‘*fp’* function in Stata statistical software (version 16.1) to incorporate longer-term time trends as fractional polynomials in all models as appropriate. We hypothesized that there would be both a step (i.e., level) and gradual (i.e., slope) change following the start of the pandemic in the number of contacts per user. For this, we included a binary coded variable in the model which represented the pandemic period (i.e., model step change), as well as an interaction term between time and intervention which models a slope change. All statistical analyses were conducted using Stata 16.

### 2.5 Ethics

The study was conducted in accordance with the Declaration of Helsinki, and approved by Ethics Committee of SWANSEA UNIVERISTY MEDICAL SCHOOL REC reference 2020-0050 on 8-10-2020.

## 3. Results

### 3.1. Routinely collected user’s demographic characteristics

There were 61221 active consenting users during the study period with a roughly similar mean number of contacts per month in the pre- and during the pandemic period (Supplementary Table 3). Consenting and non-consenting users sample distributions presented significant but very small differences in demographics with very small effect-sizes ranging v=[.06 -.07]. Males provided the highest frequency of research consent 61.1% relative to gender (X^2^(4, 107670)=542; p<.001) and those who did not state or self-reported their ethnicity with the lowest rate of consent rates 55.71% (X^2^(3, 107670)=142, p<.001) between consenters and non-consenters (Supplementary Tables SD1E and SD1G).

From the sample of consented users composing the study 80.2% were White (*N*=49070) and 74.2% female (*N*=44412). Ages ranged from 13 to 20 years of age with an average of 16.1(*SD*=1.96). From the sample extracted, 1003 participants identified as agender and 1732 as gender-fluid representing 3.4% of the total sample. Finally, almost 2% of participants did not state their ethnicity, with no difference in the level of missing by the pandemic period.

### 3.2. Change in Service access

There was evidence of a positive pre-pandemic trend in the number of contacts per user, and that there was a step-change (i.e., sudden change) in the number of contacts at the start of the pandemic (Figure 1 and Table 1).

**Figure 1.**
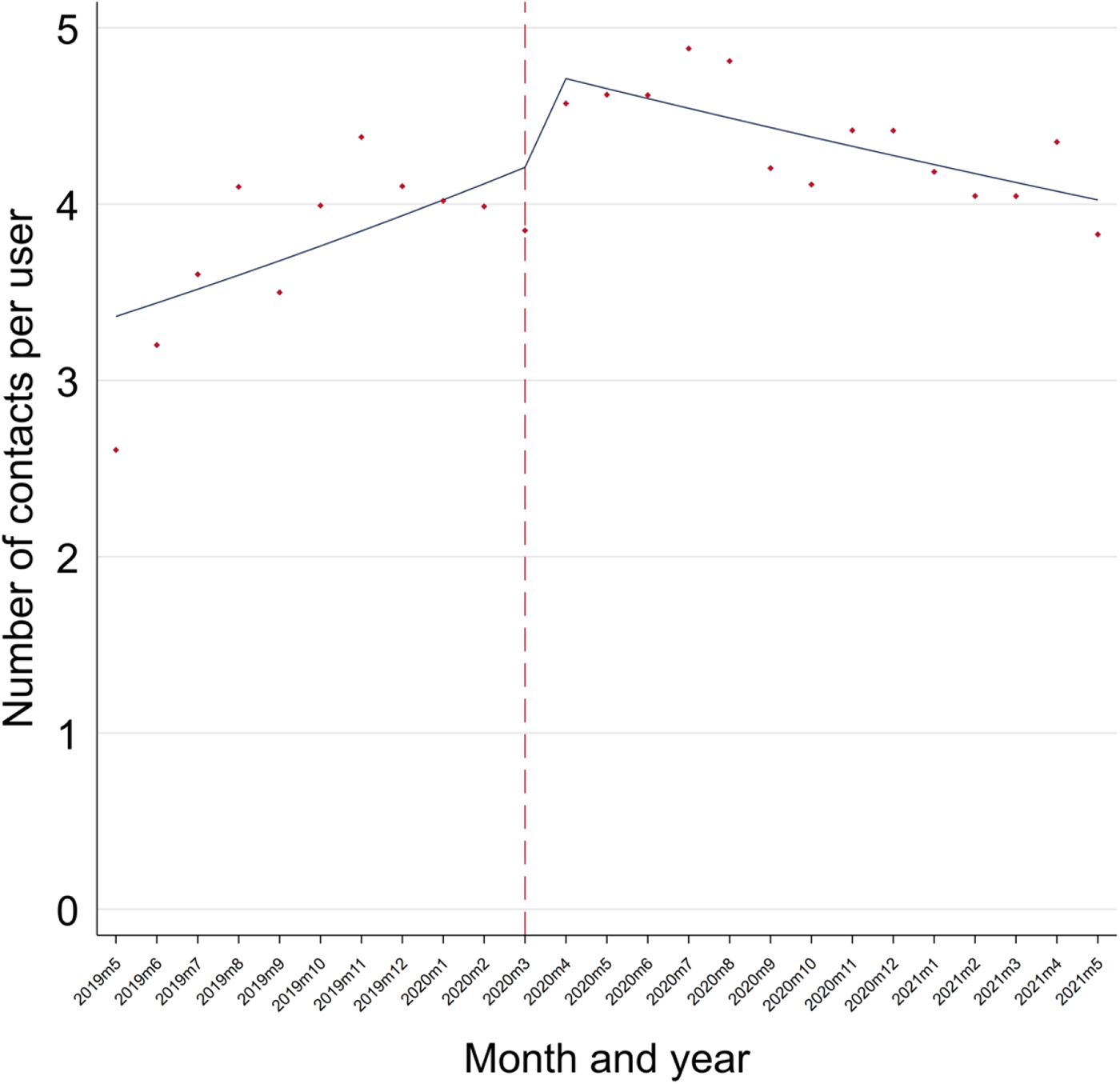
Time-series of the number of contacts per user pre- and during Covid-19 at the Digital Mental Health service.

When comparing trends in the periods, before the pandemic there was evidence of a 25% (95% CI: 9%-44%) increase rate in contacts per user per month (p=.001), this reduced to a 21% (95% CI: 4%-41%) increase per month during the pandemic (p<.001)

**Table 1.**
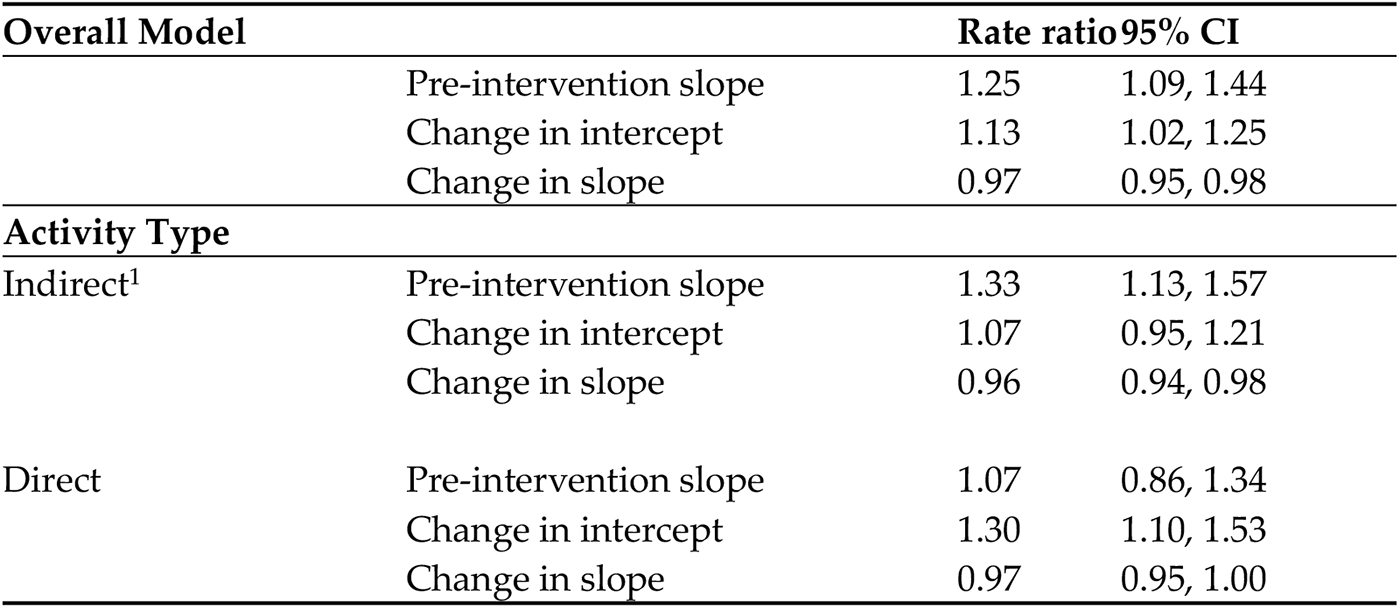
Interrupted time-series model split by type of activity in the service.

When stratified by type of activity, evidence suggests that before the pandemic there was a steady increase in the number of indirect contacts per user, with limited statistical evidence of any immediate change at the start of the pandemic (Figure 1.). There was evidence that the rate of change during the pandemic was different to the trend prior any Covid-19-related impact. Before the pandemic, there was evidence of a 33% (95% CI: 13%-57%) increase in contacts per user per month, but this rate of change reduced slightly to 28% (95%: CI 7%-54%) increase per month during the pandemic period. For direct contacts, there was no evidence of an increase in the number of contacts per user per month pre-pandemic, but evidence of a step-change increase (30%; 95% CI: 10%-53%) immediately following the start of the pandemic. There was no compelling evidence that the rate of change differed in the pandemic period compared to the period before.

### 3.3. Referral sources

There was clear evidence that the source of referrals to the service had experienced a step change during the pandemic period, with significant reductions in referrals via educational sources, and a rise in referrals via family/friend, other, care/social and internet sources (Table 3). However, there was no evidence for differences in the rate of the trends seen in relation to contacts with the digital service relating to referral sources changes in the pre- and during pandemic periods.

**Table 2.** Interrupted time-series split by presenting concerns pre- and during-pandemic.

| Presenting concerns |  | Rate Ratio | 95% CI |
| --- | --- | --- | --- |
| External | Pre-intervention slope | 0.78 | 0.54, 1.13 |
|  | Change in intercept | 1.4 | 1.05, 1.87 |
|  | Change in slope | 1 | 0.96, 1.05 |
| Mental Health | Pre-intervention slope | 0.83 | 0.56, 1.23 |
|  | Change in intercept | 1.81 | 1.36, 2.4 |
|  | Change in slope | 0.99 | 0.95, 1.04 |
| Physical health / Other | Pre-intervention slope | 0.87 | 0.48, 1.56 |
|  | Change in intercept | 1.5 | 0.96, 2.35 |
|  | Change in slope | 0.97 | 0.9, 1.04 |
| Risk / Abuse /<br>Safeguarding risk | Pre-intervention slope | 0.66 | 0.34, 1.27 |
|  | Change in intercept | 2.03 | 1.24, 3.32 |
|  | Change in slope | 1 | 0.93, 1.08 |
| Suicidality / Self-harm<br>/Maladaptive coping | Pre-intervention slope | 0.93 | 0.64, 1.36 |
|  | Change in intercept | 1.57 | 1.2, 2.06 |
|  | Change in slope | 1.02 | 0.97, 1.06 |

**Table 3.** Interrupted time-series model split by referral source self-reported in the digital service pre and during the pandemi.

| Referral Source<sup>1</sup> |  | Rate Ratio | 95% CI |
| --- | --- | --- | --- |
| Education | Pre-intervention slope | 1 | 0.88, 1.13 |
|  | Change in intercept | 0.82 | 0.71, 0.94 |
|  | Change in slope | 0.99 | 0.94, 1.04 |
| Family / Friends | Pre-intervention slope | 0.96 | 0.81, 1.13 |
|  | Change in intercept | 1.38 | 1.18, 1.61 |
|  | Change in slope | 1.01 | 0.95, 1.08 |
| Medical / Psych | Pre-intervention slope | 0.96 | 0.81, 1.15 |
|  | Change in intercept | 1.13 | 0.94, 1.34 |
|  | Change in slope | 1.02 | 0.95, 1.09 |
| Care / Social / Charity | Pre-intervention slope | 0.94 | 0.8, 1.11 |
|  | Change in intercept | 1.26 | 1.06, 1.48 |
|  | Change in slope | 0.98 | 0.92, 1.04 |
| Social media / Internet | Pre-intervention slope | 1.13 | 0.78, 1.62 |
|  | Change in intercept | 1.67 | 1.2, 2.33 |
|  | Change in slope | 0.91 | 0.8, 1.04 |
| Other | Pre-intervention slope | 0.92 | 0.69, 1.22 |
|  | Change in intercept | 1.5 | 1.14, 1.96 |
|  | Change in slope | 0.99 | 0.89, 1.1 |

<sup>1</sup> Self-reported referral source by the question "How do you hear about us?" at registration in Kooth.com

### 3.4. User’s mental health presenting concerns

The time-series modelling (Table 2) indicates that prior to the pandemic there was no evidence that the trend line either increased or decreased (i.e., relative stable rate of contacts by presenting concern).

Immediately after the pandemic, there was evidence of a step-change in the number of contacts per user for external (40% increase), mental health (81% increase), risk/abuse/safeguarding risk (100% increase), and suicidality/self-harm/maladaptive coping (57% increase). As the pandemic has progressed there was no evidence that the rate of change during the pandemic (the slope of the trend line) differs from that before the pandemic (Figure 2).

**Figure 2.**
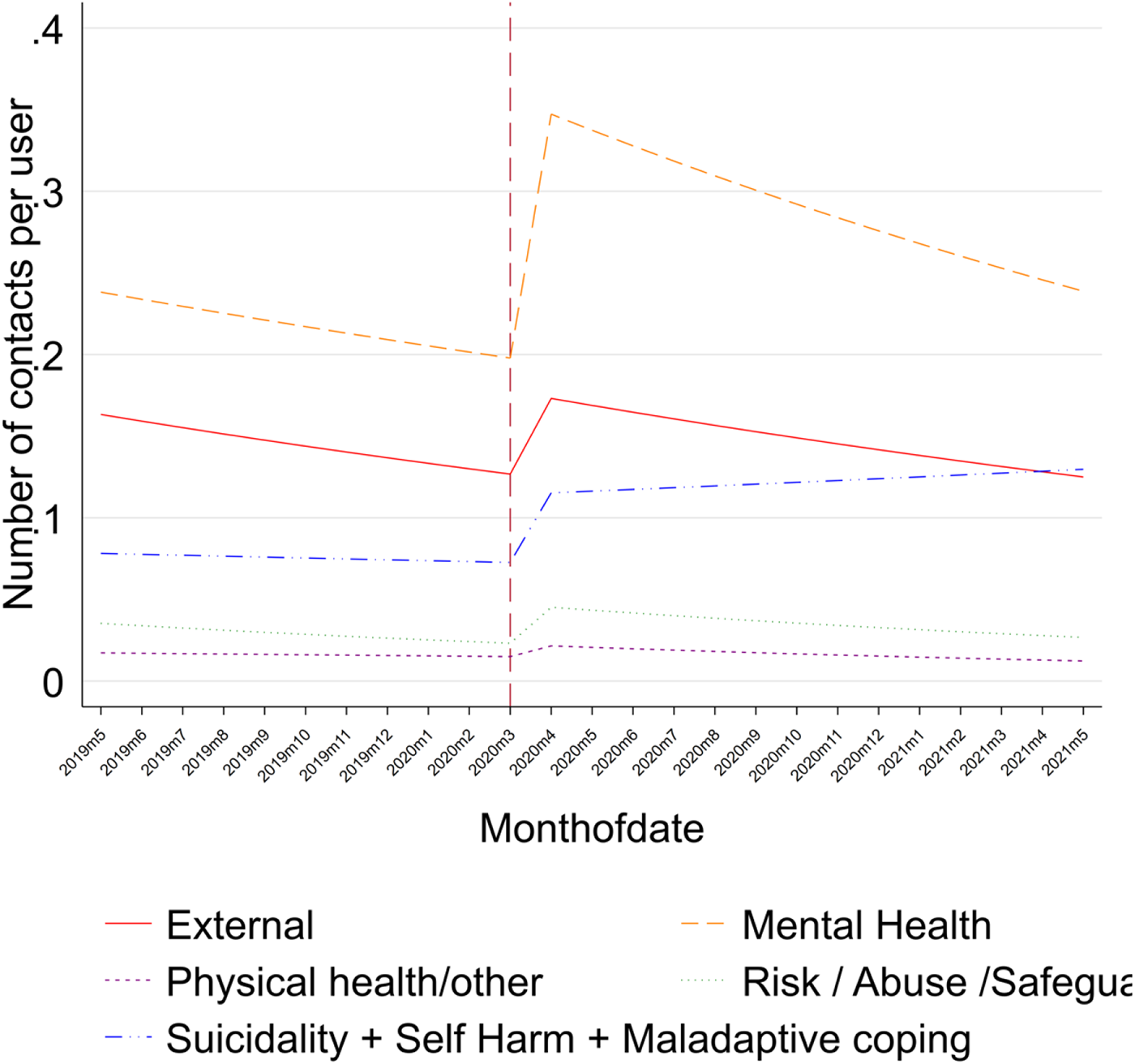
Time-series of monthly presenting concerns and contacts pre- and during Covid-19 in the digital service.

### 3.5. Ethnicity, gender, and area deprivation level service access changes

We stratified the overall contacts per user by certain demographic factors. There was evidence of a step increase in the number of contacts for Black/African/Caribbean/Black British (38% increase; 95% CI: 1%-90%) and White (14% increase; 95% CI: 2%-27%) ethnic groups. There was only evidence of a differing pandemic trend line (i.e., decreasing trend line) in the number of contacts per user for the Mixed/Multiple ethnic groups and the White majority group. When stratified by gender, we found evidence of a positive trend before the pandemic, followed by a step-change (15% increase; 95% CI: 3%-27%), and evidence that the trend line during the pandemic differed from the prior trend (i.e., decreasing trend) in females, but not other gender groups.

For the CCGs included for which we were able to derive an IMD rank linked with the public repository of deprivation information from the UK (29.4% of included CCGs had missing data for this variable). There was no evidence that the slope changed in the pandemic period compared to the pre-pandemic period for any of the deprivation quintiles (Table 5).

**Table 4.**
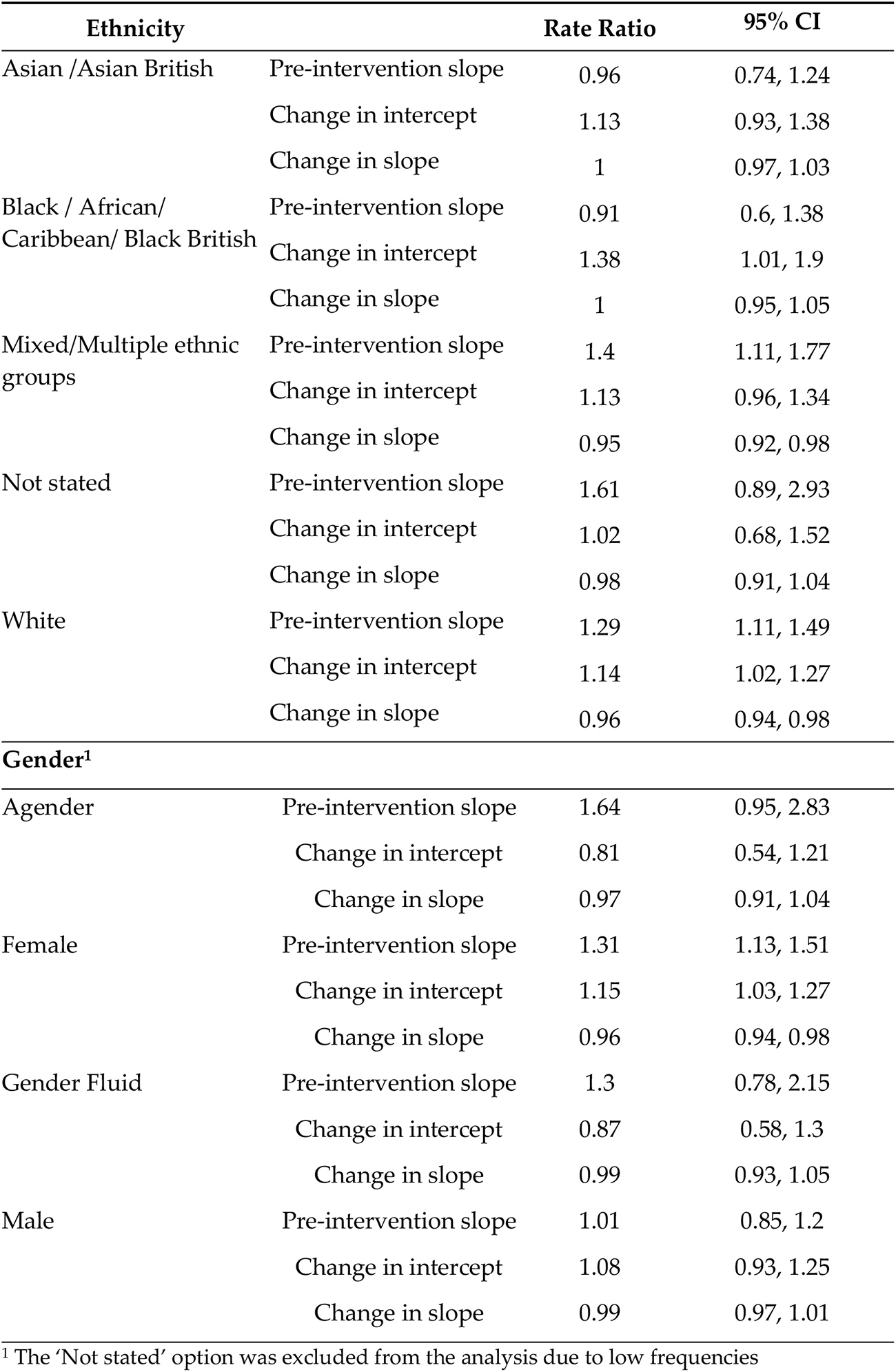
Interrupted time-series for ethnicity and gender in the digital service pre- and during-pandemic.

| Ethnicity |  | Rate Ratio | 95% CI |
| --- | --- | --- | --- |
| Asian /Asian British | Pre-intervention slope | 0.96 | 0.74, 1.24 |
|  | Change in intercept | 1.13 | 0.93, 1.38 |
|  | Change in slope | 1 | 0.97, 1.03 |
| Black / African/<br>Caribbean/ Black British | Pre-intervention slope | 0.91 | 0.6, 1.38 |
|  | Change in intercept | 1.38 | 1.01, 1.9 |
|  | Change in slope | 1 | 0.95, 1.05 |
| Mixed/Multiple ethnic<br>groups | Pre-intervention slope | 1.4 | 1.11, 1.77 |
|  | Change in intercept | 1.13 | 0.96, 1.34 |
|  | Change in slope | 0.95 | 0.92, 0.98 |
| Not stated | Pre-intervention slope | 1.61 | 0.89, 2.93 |
|  | Change in intercept | 1.02 | 0.68, 1.52 |
|  | Change in slope | 0.98 | 0.91, 1.04 |
| White | Pre-intervention slope | 1.29 | 1.11, 1.49 |
|  | Change in intercept | 1.14 | 1.02, 1.27 |
|  | Change in slope | 0.96 | 0.94, 0.98 |
| **Gender<sup>1</sup>** |  |  |  |
| Agender | Pre-intervention slope | 1.64 | 0.95, 2.83 |
|  | Change in intercept | 0.81 | 0.54, 1.21 |
|  | Change in slope | 0.97 | 0.91, 1.04 |
| Female | Pre-intervention slope | 1.31 | 1.13, 1.51 |
|  | Change in intercept | 1.15 | 1.03, 1.27 |
|  | Change in slope | 0.96 | 0.94, 0.98 |
| Gender Fluid | Pre-intervention slope | 1.3 | 0.78, 2.15 |
|  | Change in intercept | 0.87 | 0.58, 1.3 |
|  | Change in slope | 0.99 | 0.93, 1.05 |
| Male | Pre-intervention slope | 1.01 | 0.85, 1.2 |
|  | Change in intercept | 1.08 | 0.93, 1.25 |
|  | Change in slope | 0.99 | 0.97, 1.01 |

<sup>1</sup> The 'Not stated' option was excluded from the analysis due to low frequencies

**Table 5.** Interrupted Time-series model by Deprivation quartiles.

| IMD rank<br>Deprivation<sup>1</sup> | Q1 |  | Q2 |  | Q3 |  | Q4 |  | Q5 |  |
| --- | --- | --- | --- | --- | --- | --- | --- | --- | --- | --- |
|  | RR | 95% CI | RR | 95% CI | RR | 95% CI | RR | 95% CI | RR | 95% CI |
| Pre-intervention slope | 1.04 | 0.88, 1.23 | 1.61 | 0.82, 3.14 | 1.32 | 0.6, 2.87 | 1.01 | 0.99, 1.02 | 0.79 | 0.56, 1.12 |
| Change in intercept | 1.58 | 1.01, 2.47 | 1.53 | 0.91, 2.58 | 1.37 | 0.76, 2.46 | 1.65 | 0.93, 2.92 | 1.47 | 1.06, 2.04 |
| Change in slope | 1.00 | 0.99, 1.00 | 0.99 | 0.98, 1.00 | 1.00 | 0.98, 1.01 | 0.99 | 0.97, 1.01 | 1.06 | 0.93, 1.21 |

<sup>1</sup> IMD: Index of Multiple Deprivation; Q1: Most deprived – Q5: Less deprived

There was evidence that the most (58% increase; 95% CI: 1%-247%) and least (47% increase; 95% CI: 6%-204%) deprived areas both experienced a step-change increase in the number of contacts per user when Covid-19 measures commenced and the impact of the pandemic started to be experienced in the UK.

## 4. Discussion

The Covid-19 pandemic resulted in an upheaval of daily life for young people. Our results illustrate how service use and delivery in a digital mental health service immediately changed at the onset of the pandemic in March 2020 in the UK. The increase in service use observed in this study is in line with other studies based on mental health service delivery showing that the pandemic was likely to change service demands and lead to separation of care^28^. The study explored one digital mental health support service by observing changes in service access behavior before and during the pandemic with routinely collected service access data, half of the population accessing the digital mental health service consented to use their routinely collected data for research, minor distribution differences between consented and non-consented users were found in terms of demographic characteristics between samples. The observations in the consented sample reveal changes in rates of contacts observed between pre and during pandemic periods. We consider whether the crisis of the Covid-19 pandemic resulted in increased access to digital mental health support from young people and how access varied between demographic groups. Time-series analysis during this period allowed us to observe the overall access and service activity change pre-and-during Covid-19. We found that service activity changed and increase during the pandemic but changes in the referral sources and the way people access to pathways of care more generally may have impacted the increase in service access and demand, as contacts per users were higher in pre-pandemic activity when compared, our data suggest that dramatic changes in the healthcare system are likely to impact too digital ecosystems of support, even when operating anonymously and with relatively ease of access online, interoperability between services may be important in order to . We also observed changes in the presenting concerns to those likely to impact children and young people during the pandemic^29,30, 31, 32, 33^, with considerable increases on risk, self-harm and suicidal ideation, as well as other mental health related difficulties, increasing the difficulties to those more vulnerable^9^. Finally, we observed how contacts by ethnicity and area deprivation levels experienced changes in the rates of increase at the beginning of Covid-19, showing how global traumatic events like a pandemic, may be an opportunity to tackle inequality of service access to mental health support and service access online for children and young people, especially when responses to disasters are due to change these services and its use.

### 4.1. Changes in service access and activity

The beginning of the Covid-19 pandemic and the initial lockdown resulted in a sudden, negative impact on people across the UK, with young people disproportionately affected^3,4,34^. At this initial onset period (first lockdown), in terms of overall access, the number of contacts made to the digital service increased significantly, however, this was followed by a rate of engagement which was lower than what was observed before the start of the pandemic. There was evidence of trend line differences between the two periods when the increase in contact rate was compared. Other mental health services in the UK showed a reduction of service utilization during the pandemic too, along with a stable number of admissions before and during Covid-19^35,36^. One possible explanation for the change in user behaviour in digital service is that the need for support from young people peaks at the start of the pandemic. The gradual decrease in the trend of contact rate may be a consequence of most young people adjusting to the situation over time. When the psychological wellbeing at two early time points (March and May 2020) were assessed, the findings showed a deterioration in mental health symptoms among preadolescent children and a smaller deterioration in adolescents^37^. Another potential contributor to the initial increase in service use could be the reduced availability of other mental health services. A survey taken by UK mental health workers at the start of the pandemic showed that amongst community teams and treatment services, 48.4% identified not being able to depend on other services normally available as a relevant challenge^38^.

Even though there was an increase in access when the pandemic started, when looking at the overall access trend in our observational study, we see a slowing downslope. Access was seen to be increasing both before and during the pandemic, but we saw a slight drop in the rate of increase. This is interesting as we would expect, based on the prior literature an increased Mental Health need and access^39^ that there would be an exponential increase after the Covid-19 pandemic outbreak. Yet considering access more widely, we saw that referral sources dramatically changed at the start of the pandemic but the trend differences between pre and during-pandemic periods when compared were similar. This shift affected the largest referral source for the service (Schools and GPs) and a drop was observed during the pandemic.

For indirect service activity and asynchronous user contact with the service, including online forum-based activities such as discussions, comments, or posts, no evidence of an immediate change in activity at the start of the pandemic was seen. Previous literature has found community support, such as that accounted for the indirect activity in the study, to be an important factor in moderating the mental health effects of catastrophic events^40^. Sharing collective experiences can help individuals process negative events, and sharing mental health experiences online is found to support recovery and reduce stress^41,42^. Hence, it is surprising that whilst overall activity and direct contact activity increased at the start of the pandemic, indirect activity did not. We must consider that, even though the service examined here is digital, it is a digitally enabled therapy service which has trained counsellors interacting with users and moderating and monitoring the indirect and asynchronous activities users do daily. Therefore, resourcing of professionals and their tasks should be considered for the type of interaction that is requested and needed by users, as demand is likely to increase for the service in future disasters and global emergencies. Digital services service must be able to manage this demand and decide what type of resource increase will be best suited when a catastrophe takes place to meet the rising demand expected. The wider system should still consider the extra support required and importance of this digital services as part of the emergency strategy assessments to support mental health, as pathways in mainstream care and ways to access are likely to change due to emergencies like a pandemic.

### 4.2. Changes in referrals and pathways to receive digital care

The Covid-19 pandemic and associated lockdown, school and healthcare restrictions had an observable impact on changes in referrals to the digital service in the UK. Looking at the observable changes in referral sources before and during the pandemic, we can examine changing behaviour of service users, and extrapolate to the wider mental health system during the pandemic. Overall, referral sources changed significantly during the captured time-period; something that has been observed at other face-to-face mental health services during the pandemic period too^43^, other services also experienced fewer referrals during the pandemic^44,45^. The referral source for the digital service is recorded by asking the users where they ‘*heard about the service*’ when registering on the digital platform. Most of the studies observing referrals account for formal external referrals (e.g., GPs, schools) where administrative and assessment processes may be required for a patient to be accepted.

Lockdowns forced service delivery to change across the mental health provisions, and the change in referrals shows that digital services were not exempt from this impact. There was no evidence of a difference in the increase referral trend pre and during the pandemic on any source, but a spike of changes occurred at the onset of the event increasing and decreasing in some groups and illustrating referral type changes and different pathways of care to receive digital mental health support. A decrease in referrals from schools and social care services was found, whilst an increase in referrals from friends, word of mouth, primary care, and specialist mental health services was observed at the start of the pandemic. The cause for the reduction of referrals from schools was clear; pre-pandemic the digital service was frequently promoted across schools in the UK. With schools closed, this promotion was no longer possible. The increase in referrals via word of mouth and friends suggests that young people were talking more frequently online and offline about alternative care pathways for mental health support, including digital services like Kooth. The increase in specialist and primary care referrals may be an indication of integrated efforts of specialist mental health services, other professionals, and the digital service itself promoting alternatives to face-to-face services not operating as usual during Covid-19 and subsequent lockdowns, especially to those who needed an in mediate risk management action plan due their presenting concerns and current difficulties at the time of the event.

### 4.3. User’s mental health presenting concerns

Alongside looking at service use and access changes, it was important to explore the differences in presenting concerns when reaching digital support. Despite the observable presenting concerns being varied and rich, we grouped guided by the literature the most likely to be impacted by a pandemic (Supplementary Table S2). For the presenting concerns of mental health, risk, safeguarding, external difficulties, and suicide and self-harm a significant step-change increase was found in the time-series at the first lockdown. Issues of risk, abuse and, safeguarding have been a particular concern in the UK throughout the lockdown periods with typical avenues where a child at risk would be identified as being restricted, such as schools^46^ or community support. Anonymous and text-based services such as Kooth may have provided vulnerable children and young people at risk a support system replacement at a time when access to standard face-to-face support was restricted, and communication such as telephone and video calls may have been difficult in the confined environment of lockdowns^9^.

Suicide and self-harm were also presenting concerns where a sudden change was found at the start of the pandemic. As was the case for the trend in risk and safeguarding, there was not enough evidence to sustain a difference in the rate of change for contacts of users before and during the pandemic when comparing the two periods. During the period between the start of the pandemic in March 2020 and December 2020, there was no increase in child suicide rates^47^. Across ten countries, including England and Scotland, an analysis of hospital admission data showed that child and adolescent self-harm admissions increased in 2020 compared to 2019^48^. The increased contact from service users presenting with suicidal ideation and self-harms corresponds with the latter findings and suggests that though successful suicide attempts may not have increased, aborted attempts and suicidal ideation are likely to have increased during the pandemic. Previous research has shown that anxiety and depression symptoms increased amongst children and young people during the pandemic^49–51^, but is difficult to ascertain if only these two common mental health difficulties drive the increase in more specific mental health concerns for users in our study, and how those may have affected more to some than others.

### 4.4. Changes in access by sociodemographic characteristics

The pandemic was an event that impacted the UK population, though that is not to say everyone was affected equally. When looking at access to the digital service across demographic groups, there was evidence of a sudden momentary increase in contacts in those who identified as Black or White, followed by a rate of engagement which was lower in the pandemic period for White individuals, and those who identified as being from a Mixed ethnic background.

Individuals who identified as females showed a positive trend pre-pandemic, followed by a step change increase at the start of the pandemic, and then a comparative decrease in trend over the pandemic period. Other gender groups did not show significant evidence of changes. Given that females make up most of the population accessing the service, it is likely that their behaviour drove the main effects on access. A potential cause for the difference between females compared to other gender groups is the difference in sample size; females accounted for 74.2% of the service users in the sample. The limited number of other gender groups will have reduced the likelihood of consolidating the findings of such groups.

The increase in contact amongst different service users from Black ethnic backgrounds is a good sign regarding the accessibility of the service, as Black minority groups are less likely to engage in face-to-face support, such as contacting their GP about mental health^12^. Previously it has been found that CYPs from Black and Asian ethnic backgrounds are less likely to seek mental health support through voluntary referral routes^52^. This may indicate that CYPs from ethnic minorities are less likely to access early support services and thus referral sources are highly important to explore if digital support enables ethnic minorities meaningful access. However, this will raise the question of what underlying factors contribute to preferred digital engagement by the different Black communities in the UK. Our findings here correspond with other research during the pandemic, showing that ethnic minority groups were more likely to access support through helplines or online services compared to their white counterparts^53^. Our findings also suggest that there were no differences before and during the pandemic in the rate of access and contact to services, suggesting that engagements continued at a normal increase rate during the pandemic. In comparison, White service users showed a slight decrease in the rate of access during the pandemic compared to before this ethnicity the one that most commonly access Kooth in the UK and may reflect the lack of differences in access trend rates before and during the pandemic.

Referrals from word of mouth and friends increased during the pandemic. Given that mental health stigma is found in some ethnic minority communities, further analysis of differences in referral sources between different ethnic groups before and during the pandemic would provide valuable insights into whether the Covid-19 pandemic had an impact on mental health pathways for different cultural groups. A potential reason for the increased access from ethnic minorities to digital services is the anonymity provided. Anonymous online support offers the potential to reduce the perceived stigma associated with mental health-seeking, enabling CYP to access services where they might not in-person^54^. Research into causes for low mental health access from ethnic minorities has raised mental health stigma as a key contributor^55,56^, which is likely to increase the importance of anonymity amongst Black CYP.

There is an existing contrast in regards to mental health access across the areas and their socioeconomic deprivation for CYP in the UK^57^, those can affect broadly the prognosis and prevention of young people’s mental health. In our study, the sudden increase in access at the beginning of the pandemic was observed for both the most and less deprived areas from the CCG regions selected in the study. No differences were found across groups pre- and during the pandemic across deprivation quartiles. Other studies have found that whilst mental health symptoms of anxiety or depression were generally event-related and temporary ^58–60^, a tenth of the population will continue to experience enduring distress. Amongst those more at risk are people from deprived areas, minorities or with existing conditions ^61^. Previous research has highlighted the risk of the digital divide on equality of access in non-urban and rural areas ^62^, and questioned how a digital platform can be used for deprived communities where internet access is not available at home. The deprivation findings in the digital service indicate a good level of access in typically hard-to-reach populations, we also observed how access was impacted by the way the service promotion reach the individual user.

### 4.5. Study strenghts and limitations

This is an observational study of routinely collected data at a digital mental health service operating nationally in the UK, the volume of data collected is a clear strength of the study, but routinely collected data can have inconsistencies. We selected regions where there were contract stability meaning there was no change in resources during periods of interest. We also compared sample distributions between consented and non-consented users to improve the understanding of our findings. This allowed us to see differences in ethnic minorities with large volumes of data and make use of public data (deprivation) to make inferences in our observations, by observing access changes on presenting problems, platform activity and referral sources.

Despite these strengths, the study should be interpreted considering its limitations. Firstly, 50% of service users consented to their data being used. This may have impacted our findings, yet there were no large differences in the demographic distribution between the samples of consent. Secondly, the service users were primarily White or identified as Female which does not necessarily reflect the underlying population who are experiencing mental health challenges that need support. Thirdly, there are limitations in using the multiple deprivation index, as this is an area-level measure. This limits the interpretations of our findings due to its large scope in at a CCG regional level. It could be that more affluent young people in the most deprived areas accessed the service, and we are unable to disentangle this from the dataset.

### 4.6. Future research

Future studies should continue to examine access preferences for digital mental health interventions. Further attention should be given to analysing and observing user behaviour and motivational factors influencing help-seeking behaviour on these online platforms. Studies should continue exploring what is hindering access to specific groups and how digital technology and support services should be designed to promote equity of access to mental health^63^. Studies advancing in the data linkage options with different public reliable and available data repositories can give us a real understanding of access to digital mental health and help us understand what factors influence access. In turn, this can help us to understand how digital interventions can contribute to positive outcomes in mental health and respond to a global crisis^39^.

## 5. Conclusions

The Covid-19 pandemic had a significant impact on access by CYP to a digital mental health service in the UK. Activity and contact with digital web-based therapy increased in the first few months of the pandemic, but no significant differences were found between pre and during the pandemic in service activity. Whilst the cause for the decrease in access as the pandemic continued is not known, this initial increase points towards a greater need for support at the beginning of a crisis period, which in the future should be met with an increase in resources for services to help at the point of need.

Significant changes were observed in referral routes to the service, especially from educational settings. Changes were observed presenting concerns for service users before and during the pandemic, reflecting some of the immediate mental health impacts, including a significant increase in risk, abuse, and safeguarding issues for CYP but also mental health concerns, suicidality and, self-harm demonstrating the enhancing vulnerability for CYP during the pandemic. The digital service experienced similar disruption and changes in presenting problems and activities to other face-to-face healthcare settings care pathways and referrals changed because of the Covid-19 pandemic and lockdowns, the service observed changes in increasing access and service utilization by White and Black minority users, as well as the most and less deprived areas for the study.

## Supplementary Materials

see other document

## Author Contributions

“Conceptualization, S.D.O.G., D.K., A.J. and A.S.; methodology, D.K, S.D.O.G. and LS; validation, A.J. and A.S.; formal analysis, D.K.; investigation, D.K, S.D.O.G, LS, A.J. and A.S..; resources, A.S. and A.J.; data curation, S.D.O.G and D.K.; writing—original draft preparation, S.D,O.G and L.S..; writing—review and editing, D.K., S.D.O.G, L.S, L.M-C, A.J, and A.S; supervision, A.J. and A.S..; project administration, S.D.O.G.; . D.K and S.D.O.G share joint first authorship and all authors have read and agreed to the published version of the manuscript.

## Informed Consent Statement

Informed consent to share their data for research was obtained from all subjects involved in the study. Only routinely collected data from users who consented to use their information anonymously for third-party research purposes was included in the study.

## Data Availability Statement

The data presented in this study are available on request from the corresponding author and the organisation. The data are not publicly available due to privacy restrictions.

## Acknowledgments

Prof. David Gunell to help put the team together, Charlotte Mindell and Crystal Oppong to support the project and secure the internal resources at Kooth Plc. Further thanks to Kooth analyst team Baptiste Duthoit, Cristina Gascón Garcia, Ellen Howard and Tom Kayll to support data extraction. Finally, to Luke Player and Yasmin Friedman to support the project administration in the ADP trusted research environment and the wider team at Swansea University.

